# Post-COVID Syndrome. A Case Series and Comprehensive Review

**DOI:** 10.1101/2021.07.17.21260655

**Authors:** Juan-Manuel Anaya, Manuel Rojas, Martha L. Salinas, Yhojan Rodríguez, Geraldine Roa, Marcela Lozano, Mónica Rodríguez-Jiménez, Norma Montoya, Elizabeth Zapata, Post-COVID study group, Diana M Monsalve, Yeny Acosta-Ampudia, Carolina Ramírez-Santana

## Abstract

The existence of a variety of symptoms with a duration beyond the acute phase of COVID-19, is referred to as post-COVID syndrome (PCS). We aimed to report a series of patients with PCS attending a Post-COVID Unit and offer a comprehensive review on the topic. Adult patients with previously confirmed SARS-CoV-2 infection were systematically assessed through a semi-structured and validated survey. Total IgG, IgA and IgM serum antibodies to SARS-CoV-2 were evaluated by an electrochemiluminescence immunoassay. A systematic review of the literature and meta-analysis were conducted, following PRISMA guidelines. Univariate and multivariate methods were used to analyze data. Out of a total of 100 consecutive patients, 53 were women, the median of age was 49 years (IQR: 37.8 to 55.3), the median of post-COVID time after the first symptoms was 219 days (IQR: 143 to 258), and 65 patients were hospitalized during acute COVID-19. Musculoskeletal, digestive (i.e., diarrhea) and neurological symptoms including depression (by Zung scale) were the most frequent observed in PCS patients. A previous hospitalization was not associated with PCS manifestation. Arthralgia and diarrhea persisted in more than 40% of PCS patients. The median of anti-SARS-CoV-2 antibodies was 866.2 U/mL (IQR: 238.2 to 1681). Despite this variability, 98 patients were seropositive. Based on autonomic symptoms (by COMPASS 31) two clusters were obtained with different clinical characteristics. Levels of anti-SARS-CoV-2 antibodies were not different between clusters. A total of 40 articles (11,196 patients) were included in the meta-analysis. Fatigue/muscle weakness, dyspnea, pain and discomfort, anxiety/depression and impaired concentration were presented in more than 20% of patients reported. In conclusion, PCS is mainly characterized by musculoskeletal, pulmonary, digestive and neurological involvement including depression. PCS is independent of severity of acute illness and humoral response. Long-term antibody responses to SARS-CoV-2 infection and a high inter-individual variability were confirmed. Future studies should evaluate the mechanisms by which SARS-CoV-2 may cause PCS and the best therapeutic options.

**Highlights:**

- PCS is mainly characterized by musculoskeletal, pulmonary, digestive and neurological involvement including depression.
- PCS is independent of severity of acute illness and humoral immune response.
- Autonomic symptoms may help to classify patients with PCS.
- Long-term antibody responses to SARS-CoV-2 infection and a high inter-individual variability were confirmed.

## 1. Introduction

During acute infection of severe acute respiratory syndrome coronavirus 2 (SARS-CoV-2), responsible of the coronavirus disease 2019 (COVID-19), symptoms vary from mild forms to critical and more severe cases [1–3]. Symptoms in the mildest forms include dry cough, fatigue, anosmia, and fever. On the other hand, in the most severe forms, the symptoms can progress to respiratory failure requiring invasive mechanical ventilation [2,4,5]. Although most of the COVID-19 patients recover completely, without sequelae, many patients may continue experiencing COVID-19 symptoms after recovery and others may even develop new symptoms [6]. Altogether, this clinical spectrum occurring after acute infection is called post-COVID syndrome (PCS) [7]. Some authors have defined PCS as the presence of signs and symptoms after acute COVID-19 infection for more than 12 weeks [8, 9].

Among the most frequently reported PCS symptoms are fatigue, headache, attention deficit, hair loss, dyspnea, myalgia, and arthralgia [10]. However, a wide variety of symptoms have been reported within the PCS involving multiple organs and systems demanding long-term follow-up [8, 9], and even rehospitalization due to severity of PCS. In addition, most of these patients have comorbidities such as cardiovascular diseases, diabetes mellitus, obesity, cancer, and chronic kidney diseases [8, 9]. Therefore, it is crucial to understand the heterogeneity of PCS. The objective of this work was to describe the clinical and serological characteristics (i.e., antibodies anti-SARS-CoV-2) of the first 100 consecutive post-COVID patients attending a post-COVID Unit in Bogota, Colombia. In addition, we conducted a systematic review and meta-analysis on the topic.

## 2. Methods

### 2.1. Study design

A cross-sectional study was conducted from March 18^th^ to May 20^th^, 2021, at the post-COVID Unit lead by the Center for Autoimmune Diseases Research (CREA) at the Clínica del Occidente in Bogotá, Colombia. Patients older than 18 and previously confirmed SARS-CoV-2 by PCR in swab or sputum were invited to voluntary attend the post-COVID unit. The first 116 consecutive patients attending the post-COVID unit were assessed. Then, patients with history of vaccination against SARS-CoV-2 (n: 10), autoimmune diseases prior acute COVID-19 (n: 2), and patients without confirmed COVID-19 (n: 4) were excluded. A final sample size of 100 patients was included in the analyses. Of these patients, seven were asymptomatic during acute COVID-19.

Patients were systematically evaluated for post-COVID manifestations (see below), including depression and autonomic symptoms by validated scales, as well as for clinical characteristics during the acute COVID-19. All patients were tested for total anti-SARS-CoV-2 antibodies (see below for details). This study was done in compliance with Act 008430/1993 of the Ministry of Health of the Republic of Colombia, which classified it as minimal-risk research. All the patients were asked for their consent and were informed about the Colombian data protection law (1581 of 2012). The institutional review board of the Universidad del Rosario approved the study design.

### 2.2. Survey validation

A semi-structured survey was constructed based on internationally validated questionnaires that sought information during and after COVID-19 acute infection [11–15]. It was validated by a consensus of expert physicians. Once validation and approval were obtained, a pilot test was done in a group of 30 volunteers. The pilot phase allowed to identify additional questions to assess systemic compromise, to organize queries, and adapt them to guarantee its interpretability by the respondent.

### 2.3. Survey and data collection

A total of 177 questions were included in the semi-structured survey, and distributed in the following areas: identification and consent, 6; sociodemographic and epidemiological characteristics, 12; past medical history, 15; diagnosis and clinical presentation of acute COVID-19, 29; current general state of health, 4; constitutional symptoms, 4; neuropsychological (including the composite autonomic symptom score 31 [COMPASS 31] and Zung scales), 58; sense organs, 10; cardiovascular, 6; pulmonary, 8; musculoskeletal, 11; dermatological, 4; gastrointestinal, 3; and COVID-19 vaccination information, 7 (https://forms.gle/QeD96DY6NZz53hAy7). All data were collected in an electronic and secure database as described elsewhere [16]. Depression was assessed by the Zung scale. This feature was categorized as follows: Zung score < 40 absence of depression, and Zung score ≥ 40 depression [17]. Autonomic symptoms were evaluated by COMPASS 31 [14, 18].

### 2.4. Antibodies anti-SARS-CoV2

Total IgG, IgA and IgM antibodies to SARS-CoV-2 were evaluated in serum samples through the Elecsys Anti-SARS-CoV-2 electrochemiluminescence immunoassay “ECLIA” (Roche Diagnostics International AG, Rotkreuz, Switzerland). The Elecsys anti-SARS-CoV-2 S assay detects antibodies to SARS-CoV-2 Receptor Binding Domain (RBD) in a double-antigen sandwich assay format. The protocol was followed according to manufacturer instructions. Positive results by the ECLIA require a signal-to-cut-off (S/Co) value of ≥ 80 U/mL. Serum samples were initially analyzed directly without dilution, in case of results >250 U/mL, the serum sample was diluted 1:10, according to manufacturer’s recommendations. An internal validation procedure was performed, that included samples previously tested by enzyme-linked immunosorbent assay (ELISA, Euroimmun, Luebeck, Germany), and a neutralizing antibody assay (Supplementary material 1).

### 2.5. Information sources and search strategy for systematic review

A systematic review of the literature was done following the Preferred Reporting Items for Systematic Reviews and Meta-analyses (PRISMA) guideline [19]. PubMed was systematically searched for published and unpublished studies. Additional manual searches of the references cited in the articles were done. The search included articles up to May 8^th^, 2021. No restrictions were placed on study period or sample size. Other information sources such as personal communications and author’s repositories were included. Terms used for this search were: (“Post-COVID” OR “Long COVID”) AND (“COVID-19”). Articles in Spanish and English were included.

### 2.6. Eligibility criteria

Studies meeting the following criteria were included: (a) studies describing clinical manifestations after acute COVID-19, (b) studies evaluating patients in clinical settings, (c) case series, cross-sectional, case-control, cohort, or clinical trial studies were also included, (d) studies including data from national registries or unaudited databases were excluded.

### 2.7. Study selection

Study selection was done independently by three reviewers (i.e., GR, ML, JMA) who evaluated studies for eligibility in a two-step procedure. In the first phase, all identified titles and abstracts were evaluated to ensure the relationship with PCS. The potentially relevant articles were subsequently selected and evaluated again in the second phase. Here, a full-text review was done to determine whether the studies effectively reported the data about the clinical features of PCS. Retrieved articles were rejected if the eligibility criteria were not meet, and a fourth reviewer (i.e., MRJ) was consulted in cases in which the eligibility criteria were not clear.

### 2.8. Data extraction and quality assessment

Data were extracted using a standardized form to include the following variables: author, country, region, age, gender, post-COVID time, and clinical features as specified for our clinical study. A single author (i.e., GR) extracted the information, and a second reviewer (i.e., ML) verified the extracted information. Any discrepancies or missing information were resolved by consensus.

Given the discrepancies across the studies regarding the reporting of clinical manifestations, several clinical features were gathered to improve the reliability and interpretability of meta-analysis. These clinical manifestations included: impaired concentration, cognitive impairment, asthenia, dementia, polyneuropathy, heat intolerance/flushing, dizziness, headache, dysautonomia, blurry vision, xeropthalmia/sicca symptoms, nasal congestion, sneezing/coryza, palpitations, diarrhea/vomiting, dyspepsia, fatigue, arthralgia/myalgia, dyspnea, pleuritic pain, myocarditis, anxiety/depression, burning feet pain, fever, insomnia/sleep disorders. Evaluation of quality of the eligible studies was not performed. The PRISMA flowchart for systematic reviews is presented in Figure 1.

**Figure 1.**
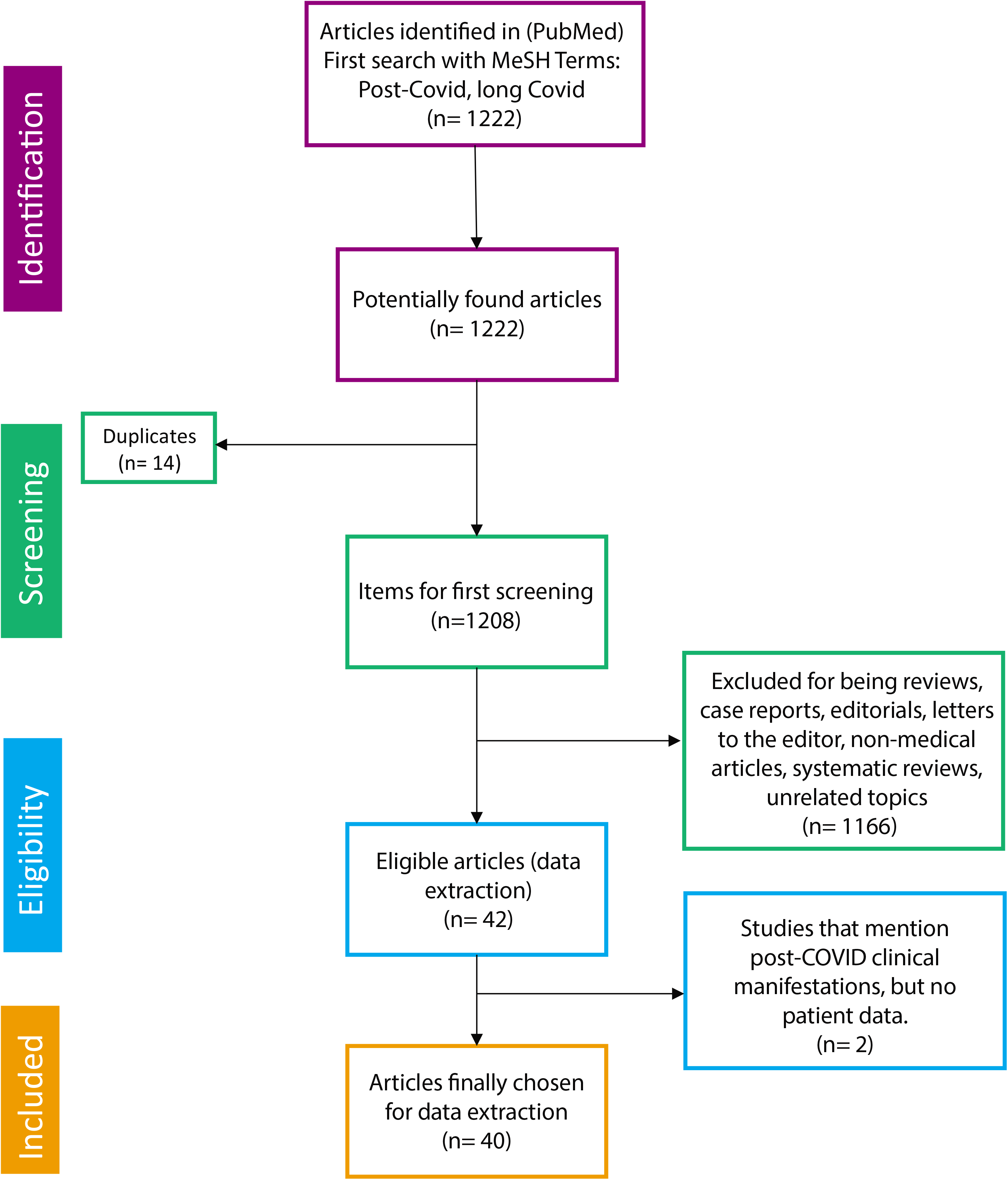
Preferred Reporting Items for Systematic Reviews and Meta-analyses (PRISMA) flow chart.

### 2.9. Statistical analysis

Univariate descriptive statistics were performed. Categorical variables were analyzed using frequencies, and quantitative continuous variables were expressed the median and interquartile range (IQR). The Kruskal-Wallis, Mann–Whitney U-test, or Fisher exact tests were used based on the results. In addition, Bonferroni correction was used for multiple testing on acute and post-COVID clinical manifestations.

To summarize the diverse information of frequencies of post-COVID manifestations, a meta-analysis approach was employed using Metafor R package (http://www.jstatsoft.org/v36/i03/). The logit transformed proportion was used to derive the weighted proportion. The overall pooled prevalence and 95% confidence intervals (CIs) were obtained using a random effect model for each clinical manifestation. Statistical heterogeneity between studies was evaluated by Cochran’s Q-statistic, as well as *Tau^2^* and *I^2^* statistics. A P value > 0.10 in Q-statistics or <50% in *I^2^* statistic indicated a lack of heterogeneity.

Next, we aimed to evaluate the likely influence of autonomic symptoms in severity of PCS, and their association with clinical phenotypes. Thus, a K-means clustering analysis based on the algorithm of Hartigan and Wong on weighted COMPASS 31 domains was conducted [20]. Shortly, the K-means method, aims to partition the points into *k* groups such that the sum of squares from points to the assigned cluster centers is minimized. For this analysis, 25 random sets were used, and a final optimal number of clusters were obtained based on the average silhouette width. In addition, patients were evaluated based on their clinical status during acute COVID-19. The significance level of the study was set to 0.05. Statistical analyses were done using R software version 4.0.2.

## 3. Results

### 3.1. General characteristics

General characteristics of patients are shown in Table 1. Out of a total of 100 patients, 53 were women, the median age was 49 years (IQR: 37.8 to 55.3), the median of post-COVID time was 219 days (IQR: 143 to 258), and 65 patients were hospitalized during the acute COVID-19. Constitutional symptoms, musculoskeletal and respiratory symptoms, ageusia and anosmia were the most frequent clinical manifestations registered during the acute illness (Table 1).

**Table 1.** General characteristics of 100 post-COVID patients.

| Variable | Post-COVID<br>n: 100 |
| --- | --- |
| **Sex** |  |
| Female | 53 (53.0%) |
| Male | 47 (47.0%) |
| **Age (Median -IQR)** | 49 (37.8 to 55.3) |
| **Body mass index (Median -IQR)** | 28.1 (25.1 to 30.4) |
| **Post-COVID time (days, Median -IQR)** | 219 days (143 to 258) |
| **Comorbidities (Prior COVID-19)** |  |
| Hypertension | 17 (17.0%) |
| Type 2 diabetes | 15 (15.0%) |
| COPD | 1 (1.0%) |
| Cancer | 1 (1.0%) |
| Dyslipidemia | 12 (12.0%) |
| Kidney disease | 3 (3.0%) |
| Hypothyroidism | 12 (12.0%) |
| Asthma | 0 (0.0%) |
| Coronary artery disease | 0 (0.0%) |
| **Clinical management** |  |
| Hospitalization | 65 (65.0%) |
| Days of hospitalization (Median -IQR) | 11 (7.5 to 16.5) |
| ICU | 24/65 (36.9%) |
| Days of ICU (Median -IQR) | 10 (6.5 to 14) |
| **Acute COVID-19 symptoms** |  |
| Fatigue | 79 (79.0%) |
| Headache | 67 (67.0%) |
| Myalgia | 66 (66.0%) |
| Arthralgia | 62 (62.0%) |
| Fever | 62 (62.0%) |
| Dry cough | 61 (61.0%) |
| Dyspnea | 59 (59.0%) |
| Ageusia | 53 (53.0%) |
| Anosmia | 51 (51.0%) |
| Pharyngitis | 48 (48.0%) |
| Diarrhea | 40 (40.0%) |
| Dizziness | 35 (35.0%) |
| Rhinorrhea | 34 (34.0%) |
| Confusion | 21 (21.0%) |
| Vomiting | 19 (19.0%) |
| Blurry vision | 18 (18.0%) |
| Impaired visual acuity | 13 (13.0%) |
| Edema | 10 (10.0%) |
| **Post-COVID manifestations** |  |
| Arthralgia | 65 (65.0%) |
| Back pain | 55 (55.0%) |
| Arms/legs heaviness | 47 (47.0%) |
| Weakness | 46 (46.0%) |
| Diarrhea | 46 (46.0%) |
| Headache | 45 (45.0%) |
| Myalgia | 42 (42.0%) |
| Body pain | 40 (40.0%) |
| Paresthesia | 38 (38.0%) |
| Attention disorders | 36 (36.0%) |
| Memory disorders | 36 (36.0%) |
| Fatigue | 34 (34%) |
| Depression | 32 (32.0%) |
| Dizziness | 31 (31.0%) |
| Chest tightness | 30 (30.0%) |
| Tachycardia | 29 (29.0%) |
| Chest pain | 29 (29.0%) |
| Eye pain | 27 (27.0%) |
| Anorexia | 23 (23.0%) |
| Chills | 19 (19.0%) |
| Weight loss | 20 (20.0%) |
| Exanthema | 26 (26.0%) |
| Blisters | 8 (8.0%) |
| Skin sensitivity | 15 (15.0%) |
| Sarcopenia | 28 (28.0%) |
| Palpitations | 27 (27.0%) |
| Xerostomia | 26 (26.0%) |
| Abdominal pain | 24 (24.0%) |
| Dyspnea | 24 (24.0%) |
| Tinnitus | 23 (23.0%) |
| Xerophthalmia | 23 (23.0%) |
| Vision disorders | 21 (21.0%) |
| Pharyngitis | 19 (19.0%) |
| Rhinorrhea | 19 (19.0%) |
| Sickness | 19 (19.0%) |
| Dry cough | 17 (17.0%) |
| Inability to walk | 16 (16.0%) |
| Pleurisy | 16 (16.0%) |
| Ageusia | 15 (15.0%) |
| Alopecia | 13 (13.0%) |
| Edema | 12 (12.0%) |
| Anosmia | 11 (11.0%) |
| Wet cough | 10 (10.0%) |
| Erectile dysfunction | 3 (3.0%) |
| Tooth Loss | 2 (2.0%) |
| New onset AD | 2 (2.0%) |
| Hemoptysis | 1 (1.0%) |
| Seizures | 1 (1.0%) |
| Insomnia | 1 (1.0%) |
| **COMPASS 31 score (Median -IQR)** | **9 (4 to 17)** |
| **Zung score (Median -IQR)** | **35.5 (30 to 41.5)** |
| **Thyroid stimulating hormone (μIU /mL) (Median -IQR)** | **2.2 (1.5 to 3.3)** |
| **Total anti-SARS-CoV-2 antibodies (U/mL) (Median -IQR)** | **866.2 (238.2 to 1681)** |

IQR: Interquartile range; ICU: Intensive care unit; COPD: Chronic pulmonary obstructive disease; AD: Autoimmune disease; COMPASS 31: Composite Autonomic Symptom Score 31; COVID-19: Coronavirus disease 2019; SARS-CoV-2: Severe acute respiratory syndrome coronavirus 2; Thyroid stimulating hormone, by electrochemoluminescence using the following thresholds: 0.3–4.5 μIU /mL.

### 3.2. Post-COVID manifestations

Musculoskeletal, digestive (i.e., diarrhea) and neurological symptoms including depression (35%) were the most frequent observed during PCS (Table 1 – Figure 2 A-B). One-third of PCS patients present with at least one musculoskeletal, respiratory, gastrointestinal and neurological symptoms simultaneously (Figure 2C). Interestingly, there were a reduction in the frequency of some acute symptoms reported by patients. However, arthralgia and diarrhea persisted in more than 40% of the patients during PCS (Figure 2D).

**Figure 2.**
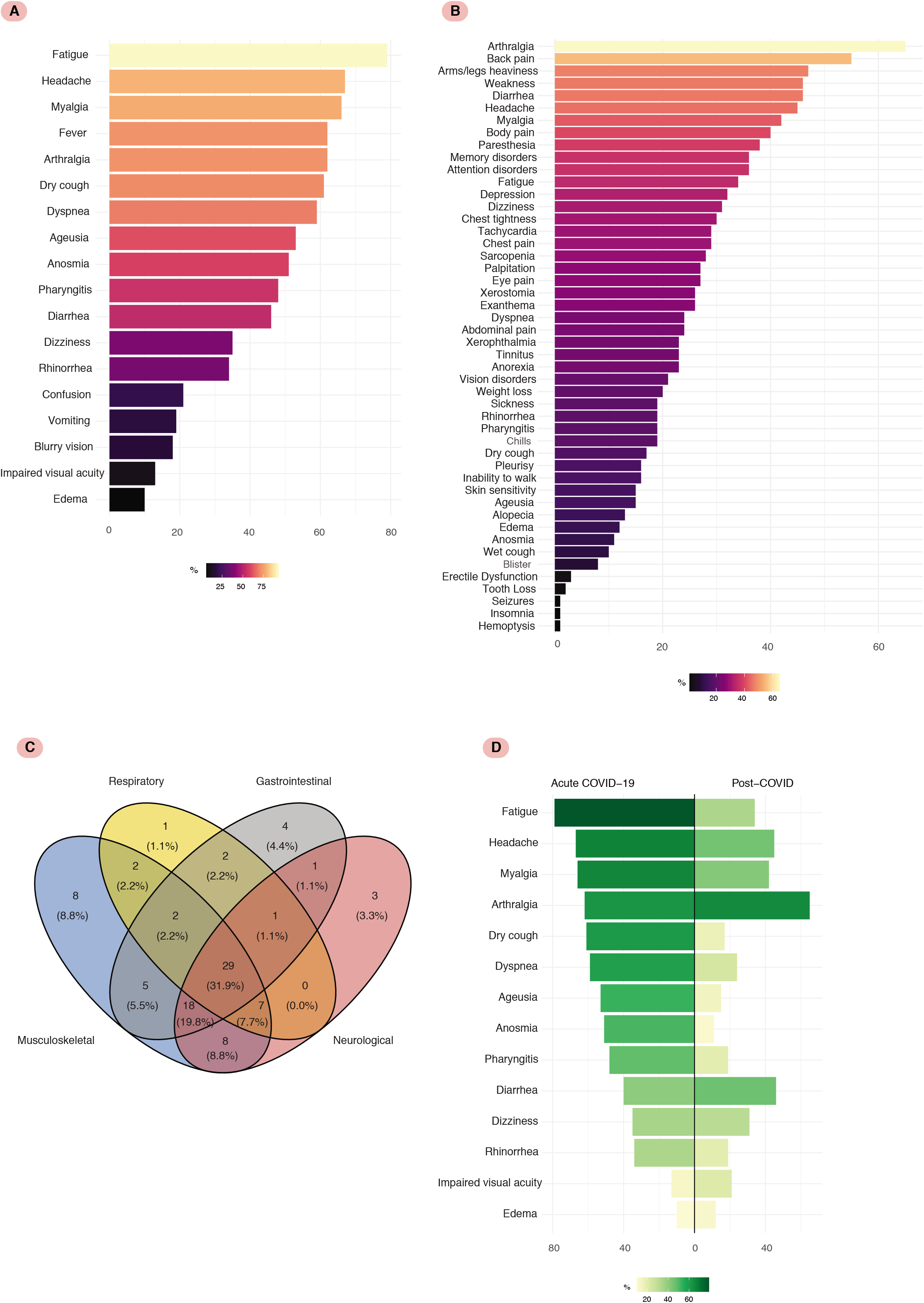
Acute and post-COVID symptoms. **A.** Frequency bar plot for clinical manifestations on acute COVID-19. **B.** Frequency bar plot for post-COVID clinical manifestations. Frequency of depression was estimated by Zung scale. **C**. Venn diagram with the superposition of the main PCS symptoms. Analysis included 91 patients, because 9 out of 100 patients did not exhibit any of the four main symptoms. **D.** Mirrored bar plot for symptoms on acute COVID-19 and post-COVID syndrome.

A median of anti-SARS-CoV-2 antibodies of 866.2 U/mL (IQR: 238.2 to 1681) was observed, indicating large inter-individual variability (Table 1). Despite this variability, almost all patients (98.0%) presented positivity for anti-SARS-CoV-2 antibodies (value of ≥0.80 U/mL, Figure 3), of whom 88 patients disclosed titles above 151.4 U/mL.

**Figure 3.**
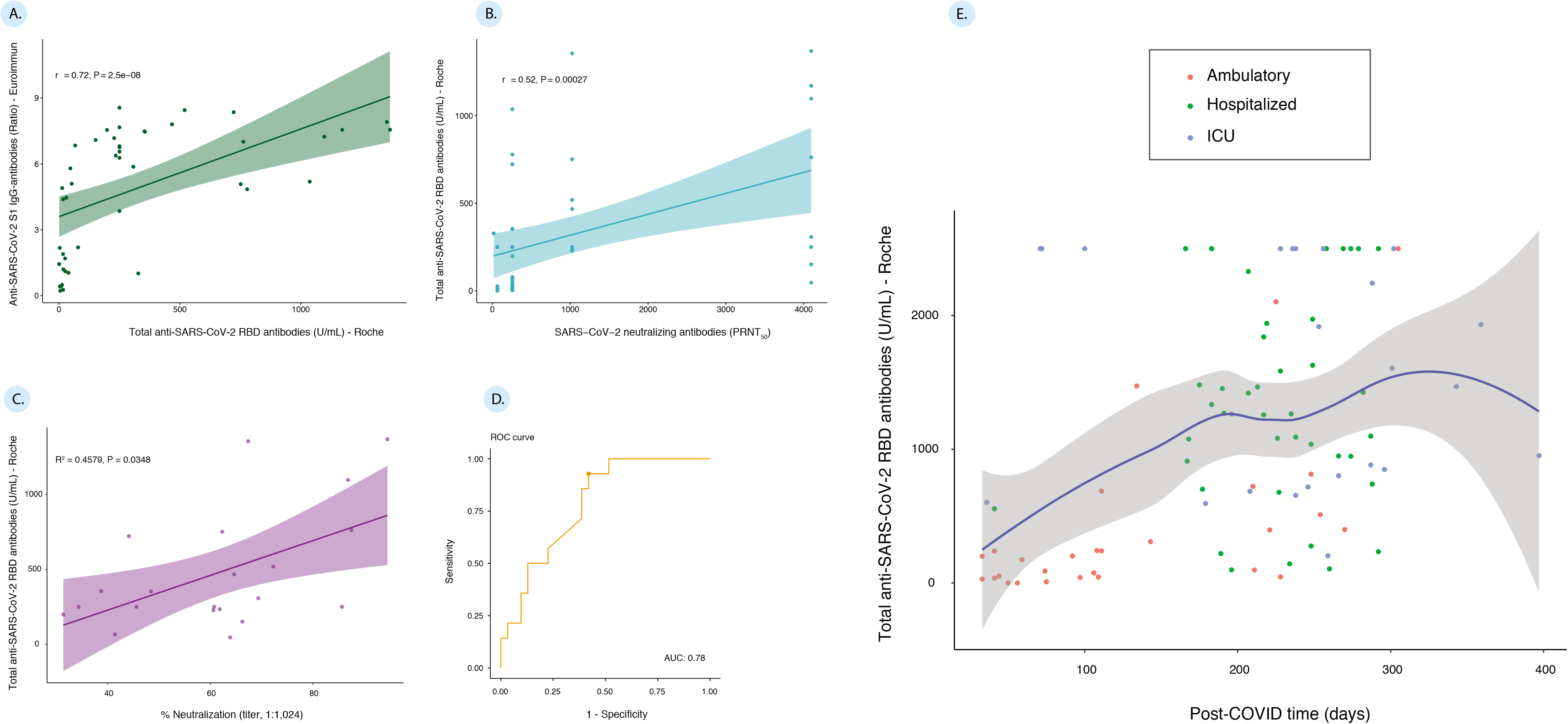
Anti RBD SARS-CoV-2 antibodies (see supplementary material 1 for details). **A.** Correlation between IgG anti-SARS-CoV-2 by ELISA (Euroimmun) and total anti-SARS-CoV-2 antibodies by ECLIA (Roche)**. B.** Correlation between neutralizing anti-SARS-CoV-2 antibodies (PRNT_50_) and total anti-SARS-CoV-2 antibodies by ECLIA (Roche). **C.** Linear model for neutralizing antibodies based on ECLIA. **D.** ROC curve for the estimated cut-off value. The AUC represents the discrimination capacity of this threshold. **E.** Scatter plot for total anti-SARS-CoV-2 antibodies (by ECLIA, Roche) and post-COVID time. Line represents the locally estimated scatterplot smoothing with 95% confidence intervals.

Evaluation of PCS based on severity of acute illness showed that hospitalized and critically ill patients were older, more likely to exhibit elevated body mass index, higher frequency of hypertension, fatigue, and fever on admission that ambulatory patients (Table 2). Noteworthy, during the PCS a previous hospitalization was not associated with any clinical manifestation. The levels of antibodies anti-SARS-CoV-2 were lower in patients who were not previously hospitalized (Table 2). However, ambulatory patients were evaluated in earlier phases of PCS than hospitalized and critically ill patients (Kruskal-Wallis test, P<0.0001).

**Table 2.** General characteristics of post-COVID patients based on severity of acute disease.

| Variable | Ambulatory<br>n: 35 | Severe disease<br>n: 41 | Critical disease<br>n: 24 | P value |
| --- | --- | --- | --- | --- |
| **Sex** |  |  |  | <0.0001 |
| Female | 30 (85.7%) | 15 (36.6%) | 8 (33.3%) |  |
| Male | 5 (14.3%) | 26 (63.4%) | 16 (66.7%) |  |
| **Age (Median -IQR)** | 41 (95.5 to 49.5) | 51 (44 to 55) | 52.5 (41.3 to 59.5) | 0.0020 |
| **Body mass index (Median -IQR)** | 25.4 (23.6 to 28.4) | 28.8 (26 to 32) | 28.3 (25.7 to 30.3) | 0.0006 |
| **Post-COVID time (days, Median -IQR)** | 108.5 (58.3 to 213.5) | 277 (190 to 260) | 250 (205 to 290) | <0.0001 |
| **Comorbidities (Prior COVID-19)** |  |  |  |  |
| Hypertension | 2 (5.7%) | 8 (19.5%) | 7 (29.2%) | 0.0361 |
| Type 2 diabetes | 2 (5.7%) | 7 (17.1%) | 6 (25.0%) | 0.0943 |
| COPD | 0 (0.0%) | 1 (2.4%) | 0 (0.0%) | 1.0000 |
| Cancer | 1 (2.9%) | 0 (0.0%) | 0 (0.0%) | 0.5900 |
| Dyslipidemia | 1 (2.9%) | 6 (14.6%) | 5 (20.8%) | 0.0733 |
| Kidney disease | 1 (2.9%) | 1 (2.4%) | 1 (4.2%) | 1.0000 |
| Hypothyroidism | 2 (5.7%) | 8 (19.5%) | 2 (8.3%) | 0.1824 |
| Asthma | 0 (0.0%) | 0 (0.0%) | 0 (0.0%) | - |
| Coronary artery disease | 0 (0.0%) | 0 (0.0%) | 0 (0.0%) | - |
| **Acute COVID-19 symptoms** |  |  |  |  |
| Anosmia | 20 (57.1%) | 22 (53.7%) | 9 (37.5%) | 0.3156 |
| Ageusia | 22 (62.9%) | 22 (53.7%) | 9 (37.5%) | 0.1764 |
| Fatigue | 19 (54.3%) | 38 (92.7%) | 22 (91.7%) | <0.0001 |
| Headache | 25 (71.4%) | 28 (68.3%) | 14 (58.3%) | 0.5896 |
| Dry cough | 16 (45.7%) | 27 (65.9%) | 18 (75.0%) | 0.0580 |
| Rhinorrhea | 12 (34.3%) | 15 (36.6%) | 7 (29.2%) | 0.8301 |
| Pharyngitis | 17 (48.6%) | 20 (48.8%) | 11 (45.8%) | 1.0000 |
| Dizziness | 11 (31.4%) | 14 (34.1%) | 10 (41.7%) | 0.7186 |
| Confusion | 5 (14.3%) | 7 (17.1%) | 9 (37.5%) | 0.0916 |
| Impaired visual acuity | 3 (8.6%) | 8 (19.5%) | 2 (8.3%) | 0.3112 |
| Blurry vision | 5 (14.3%) | 11 (26.8%) | 2 (8.3%) | 0.1586 |
| Dyspnea | 15 (42.9%) | 27 (65.9%) | 17 (70.8%) | 0.0599 |
| Diarrhea | 9 (25.7%) | 23 (56.1%) | 8 (33.3%) | 0.0203 |
| Vomiting | 4 (11.4%) | 12 (29.3%) | 3 (12.5%) | 0.1161 |
| Myalgia | 20 (57.1%) | 32 (78.0%) | 14 (58.3%) | 0.1071 |
| Arthralgia | 21 (60.0%) | 27 (65.9%) | 14 (58.3%) | 0.7796 |
| Edema | 5 (14.3%) | 4 (9.8%) | 1 (4.2%) | 0.5328 |
| Fever | 13 (37.1%) | 30 (73.2%) | 19 (79.2%) | 0.0010 |
| **Post-COVID symptoms** |  |  |  |  |
| Depression | 13 (37.1%) | 8 (19.5%) | 11 (45.8%) | 0.0661 |
| New onset AD | 2 (5.7%) | 0 (0.0%) | 0 (0.0%) | 0.1760 |
| Anorexia | 9 (25.7%) | 8 (19.5%) | 6 (25.0%) | 0.8272 |
| Chills | 7 (20.0%) | 8 (19.5%) | 4 (16.7%) | 1.0000 |
| Weight Loss | 8 (22.9%) | 6 (14.6%) | 6 (25.0%) | 0.5092 |
| Exanthema | 9 (25.7%) | 12 (29.3%) | 5 (20.8%) | 0.7705 |
| Blisters | 4 (11.4%) | 3 (7.3%) | 1 (4.2%) | 0.7217 |
| Skin sensitivity | 8 (22.9%) | 7 (17.1%) | 0 (0.0%) | 0.0347 |
| Sarcopenia | 6 (17.1%) | 11 (26.8%) | 11 (45.8%) | 0.0601 |
| Weakness | 14 (40.0%) | 15 (36.6%) | 17 (70.8%) | 0.0221 |
| Myalgia | 14 (40.0%) | 17 (41.5%) | 11 (45.8%) | 0.9333 |
| Back pain | 15 (42.9%) | 24 (58.5%) | 16 (66.7%) | 0.1752 |
| Arms/legs heaviness | 12 (34.3%) | 22 (53.7%) | 13 (54.2%) | 0.1825 |
| Inability to walk | 3 (8.6%) | 7 (17.1%) | 6 (25.0%) | 0.2323 |
| Body pain | 10 (28.6%) | 17 (41.5%) | 13 (54.2%) | 0.1420 |
| Arthralgia | 20 (57.1%) | 28 (68.3%) | 17 (70.8%) | 0.4965 |
| Fatigue | 9 (25.7%) | 15 (36.6%) | 10 (41.7%) | 0.4078 |
| Palpitations | 9 (25.7%) | 11 (26.8%) | 7 (29.2%) | 0.9593 |
| Xerostomia | 9 (25.7%) | 8 (19.5%) | 9 (37.5%) | 0.2800 |
| Chest tightness | 9 (25.7%) | 14 (34.1%) | 7 (29.2%) | 0.6980 |
| Tachycardia | 10 (28.6%) | 12 (29.3%) | 7 (29.2%) | 1.0000 |
| Edema | 5 (14.3%) | 6 (14.6%) | 1 (4.2%) | 0.4584 |
| Chest pain | 11 (31.4%) | 11 (26.8%) | 7 (29.2%) | 0.9596 |
| Hemoptysis | 1 (2.9%) | 0 (0.0%) | 0 (0.0%) | 0.5900 |
| Dyspnea | 11 (31.4%) | 8 (19.5%) | 5 (20.8%) | 0.4587 |
| Wet cough | 3 (8.6%) | 4 (9.8%) | 3 (12.5%) | 0.8373 |
| Dry cough | 4 (11.4%) | 10 (24.4%) | 3 (12.5%) | 0.3186 |
| Pleurisy | 7 (20.0%) | 6 (14.6%) | 3 (12.5%) | 0.7371 |
| Rhinorrhea | 3 (8.6%) | 9 (22.0%) | 7 (29.2%) | 0.1036 |
| Sickness | 7 (20.0%) | 10 (24.4%) | 2 (8.3%) | 0.2869 |
| Abdominal pain | 8 (22.9%) | 11 (26.8%) | 5 (20.8%) | 0.8716 |
| Paresthesia | 11 (31.4%) | 16 (39.0%) | 11 (45.8%) | 0.5271 |
| Attention disorders | 13 (37.1%) | 9 (22.0%) | 14 (58.3%) | 0.0140 |
| Memory disorders | 12 (34.3%) | 9 (22.0%) | 15 (62.5%) | 0.0047 |
| Seizures | 1 (2.9%) | 0 (0.0%) | 0 (0.0%) | 0.5900 |
| Headache | 15 (42.9%) | 18 (43.9%) | 12 (50.0%) | 0.8734 |
| Dizziness | 11 (31.4%) | 12 (29.3%) | 8 (33.3%) | 0.9616 |
| Xerophthalmia | 6 (17.1%) | 11 (26.8%) | 6 (25.0%) | 0.5947 |
| Tooth Loss | 1 (2.9%) | 1 (2.4%) | 0 (0.0%) | 1.0000 |
| Tinnitus | 10 (28.6%) | 7 (17.1%) | 6 (25.0%) | 0.4914 |
| Anosmia | 7 (20.0%) | 3 (7.3%) | 1 (4.2%) | 0.1589 |
| Ageusia | 10 (28.6%) | 3 (7.3%) | 2 (8.3%) | 0.0368 |
| Vision disorders | 4 (11.4%) | 10 (24.4%) | 7 (29.2%) | 0.1985 |
| Eye pain | 7 (20.0%) | 13 (31.7%) | 7 (29.2%) | 0.5483 |
| Pharyngitis | 4 (11.4%) | 7 (17.1%) | 8 (33.3%) | 0.1227 |
| Alopecia | 5 (14.3%) | 5 (12.2%) | 3 (12.5%) | 1.0000 |
| Erectile dysfunction | 0 (0.0%) | 0 (0.0%) | 3 (12.5%) | 0.0125 |
| Insomnia | 1 (2.9%) | 0 (0.0%) | 0 (0.0%) | 0.5900 |
| **COMPASS-31 score (Median -IQR)** | 7 (2.5 to 16) | 9 (5 to 17) | 13 (6.5 to 17.5) | 0.3221 |
| **Zung score (Median -IQR)** | 36 (29.5 to 45) | 32 (29 to 38) | 38.5 (32.8 to 44.3) | 0.1506 |
| **Thyroid stimulating hormone (μIU /mL) (Median -IQR)** | 1.90 (1.1 to 2.5) | 2.66 (2.0 to 4.0) | 2.06 (1.3 to 3.5) | 0.0130 |
| **Total anti-SARS-CoV-2 antibodies (U/mL) (Median -IQR)<sup>b</sup>** | 200.2 (52.1 to 398.2) | 1270 (911.5 to 1941) | 1537.5 (780.9 to 2500) | <0.000* |

IQR: Interquartile range; ICU: Intensive care unit; COPD: Chronic pulmonary obstructive disease; AD: Autoimmune disease; COMPASS 31: Composite Autonomic Symptom Score 31; COVID-19: Coronavirus disease 2019; SARS-CoV-2: Severe acute respiratory syndrome coronavirus 2; Thyroid stimulating hormone, by electroquimioluminescence using the following thresholds: 0.3–4.5 μIU /mL.
<sup>a</sup> P values for categorical variables were obtained by Fisher's exact test. Quantitative variables were analyzed by Kruskal-Wallis test.
<sup>b</sup> A linear regression model including total SARS-CoV-2 antibodies as dependent variable was conducted. Sex, age, body mass index, clinical management, and post-COVID time were included as covariates. Linear model confirmed that post-COVID time was a confounding factor for the difference of total SARS-CoV-2 antibodies among groups ( $\beta = 4.36$ , $P = 0.0126$ ). In addition, significant interactions for ICU management confirmed this result ( $\beta = -5.65$ , $P = 0.0221$ ).
\* Statistically significant after Bonferroni correction. P value threshold for acute COVID-19 symptoms: $0.05/18 = 0.0028$ ; P value threshold for post-COVID symptoms: $0.05/47 = 0.0011$ .

### 3.3. Post-COVID systematic review

Initially, 1222 records were found through the database search. After duplicate studies were excluded, a total of 1208 studies were obtained. After the first records were screened by title and abstract, 42 articles were fully assessed for eligibility. Of these, 2 articles were excluded, since they reported PCS clinical manifestations but not patient data. This procedure left 40 articles (11,196 patients) that fulfilled the inclusion criteria, and they were included in the quantitative and qualitative synthesis (Figure 1 and Supplementary Material 2)

### 3.4. Studies characteristics

Out of 40 studies included in the systematic review [7,10,21–58], 30 were cohort studies, 5 cross-sectional, 3 case series, and 2 case-control studies. No clinical trials were included in the systematic review and meta-analysis.

### 3.5. Post-COVID meta-analysis

Pooled prevalence of PCS manifestations is summarized in Table 3. Fatigue/muscle weakness, dyspnea, pain and discomfort, anxiety/depression and impaired concentration were presented in more than 20% of patients included in the meta-analysis (Figure 4). These symptoms were reported in more than five manuscripts in the literature review. In addition, high methodological and statistical heterogeneity was found in this analysis, accounting for most of the imprecision detected by the Q and *I^2^* statistics.

**Figure 4.**
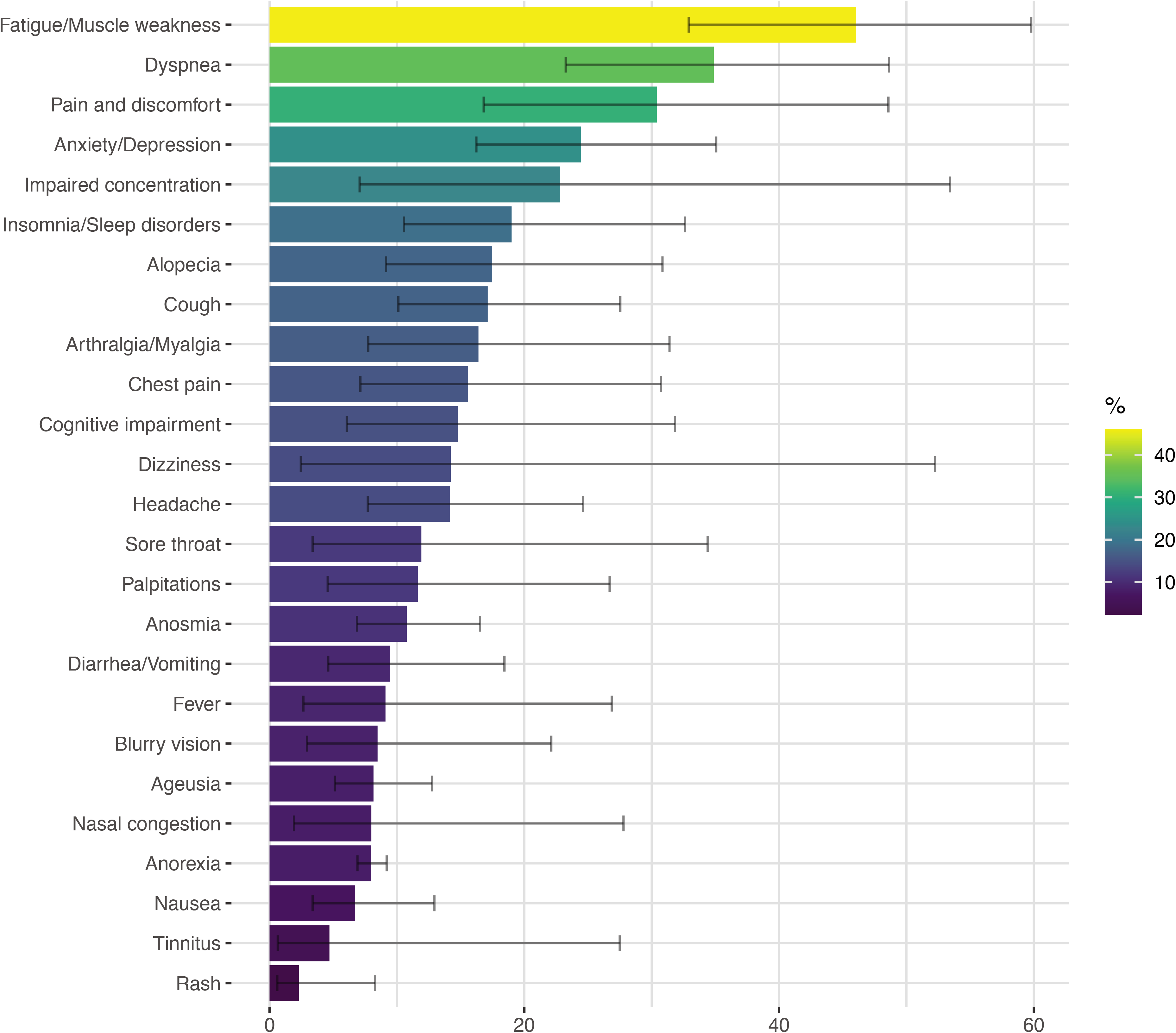
Frequency bar plot for pooled prevalence of post-COVID manifestations in meta-analysis. Error bar represents the estimated 95% confidence interval. Only those clinical manifestations reported in more than 4 articles were included in this graph.

**Table 3.** Clinical features of post-COVID-19 patients (systematic review and meta-analysis).

| Signs and Symptoms | Number of articles <sup>a</sup> | Cases/COVID-19 (% - mean, 95% CI) <sup>b</sup> | Q ($\tau^2$, $I^2$) |
| --- | --- | --- | --- |
| **Demographics** |  |  |  |
| Male | 40 | 5087/11196 (50.0%, 44.5 to 55.6) | < 0.01 (0.5, 96.3) |
| Age (years) | 34 | 10081 (50.2 years, 47.0 to 53.5) | < 0.01 (90.0, 99.4%) |
| Time to follow-up (days) | 7 | 2849 (105.9 days, 89.1 to 122.7) | < 0.01 (512.1, 99.8%) |
| **Constitutional symptoms** |  |  |  |
| Fever | 8 | 269/3064 (9.1%, 2.7 to 26.9) | < 0.01 (3.3, 98.1%) |
| Anorexia | 5 | 173/2183 (8.0%, 6.9 to 9.2) | 0.77 (0, 0.0%) |
| Weight loss | 3 | 82/2414 (4.1%, 2.8 to 5.9) | 0.01 (0.1, 41.5%) |
| Night sweats | 3 | 49/774 (5.10%, 0.8 to 26.6) | < 0.01 (2.7, 96.4%) |
| Asthenia | 2 | 49/126 (20.9%, 2.1 to 76.8) | 0.01 (2.9, 83.9%) |
| Heat intolerance/Flushing | 2 | 276/2140 (10.0%, 3.6 to 25.1) | 0.18 (0.4, 43.5%) |
| **Neurological** |  |  |  |
| Headache | 16 | 1365/7018 (14.2%, 7.7 to 24.6) | < 0.01 (1.8, 98.5%) |
| Cognitive impairment | 14 | 464/2991 (14.8%, 6.1 to 31.8) | < 0.01 (3.4, 98.4%) |
| Dizziness | 6 | 722/4912 (14.2%, 2.5 to 52.3) | < 0.01 (5.4, 99.5%) |
| Dementia | 3 | 89/405 (12.4%, 4.3 to 30.9) | < 0.01 (0.8, 82.0%) |
| Polyneuropathy | 3 | 5/69 (8.3%, 3.5 to 18.5) | 0.5 (0, 0.0%) |
| Dysautonomia | 2 | 11/58 (16.9%, 4.0 to 49.8) | 0.1 (1.0, 73.8%) |
| Inability to walk | 2 | 105/359 (26.3%, 9.4 to 55) | < 0.01 (0.8, 95.7%) |
| Stroke | 2 | 9/445 (1.8%, 0.5 to 6.6) | 0.16 (0.6, 49.9%) |
| Anomic aphasia | 2 | 7/49 (14.9%, 3.2 to 48.5) | 0.06 (1.1, 72.5%) |
| Tremor | 2 | 7/305 (3.8%, 0.2 to 48.9) | < 0.01 (5.0, 93.0%) |
| Syncopal | 1 | 3/27 (11.1%, 3.6 to 29.3) | NA |
| Restless leg syndrome | 1 | 2/355 (0.56%, 0.1 to 2.2) | NA |
| Ataxia | 1 | 1/48 (2.1%, 0.3 to 13.4) | NA |
| Orthostatic headache | 1 | 6/27 (22.2%, 10.3 to 41.5) | NA |
| Paresthesia | 1 | 13/49 (26.5%, 16.1 to 40.5) | NA |
| Delirium | 1 | 2/27 (7.4%, 1.9 to 25.3) | NA |
| **Ophthalmological** |  |  |  |
| Blurry vision | 4 | 86/746 (8.5%, 2.9 to 22.1) | < 0.01 (1.1, 93.5%) |
| Xerophthalmia/Sicca symptoms | 3 | 42/171 (14.1%, 2.7 to 49.1) | < 0.01 (2.1, 90.4%) |
| Impaired visual acuity | 2 | 41/802 (10.0%, 1.0 to 56.3) | < 0.01 (3.1, 98.0%) |
| Conjunctivitis | 1 | 2/102 (2.0%, 0.5 to 7.5) | NA |
| Eye tearing | 1 | 2/64 (3.1%, 0.8 to 11.7) | NA |
| Macular detachment | 1 | 55/87 (63.2%, 52.6 to 72.7) | NA |
| Ocular burning sensation | 1 | 15/64 (23.4%, 14.7 to 35.3) | NA |
| Red eye | 1 | 2/64 (3.1%, 0.8 to 11.7) | NA |
| Retinal cotton wool spots | 1 | 9/70 (12.9%, 6.8 to 22.9) | NA |
| Vitreous fibrillary degeneration | 1 | 10/70 (14.3%, 7.9 to 24.6) | NA |
| **Otorhinolaryngological** |  |  |  |
| Ageusia | 13 | 707/6628 (8.2%, 5.1 to 12.8) | < 0.01 (0.7, 96.4%) |
| Anosmia | 17 | 926/7302 (10.8%, 6.9 to 16.5) | < 0.01 (1.0, 97.4%) |
| Sore throat | 6 | 781/4504 (11.9%, 3.4 to 34.4) | < 0.01 (2.8, 99.2%) |
| Nasal congestion | 5 | 490/3373 (8.0%, 1.9 to 27.8) | < 0.01 (2.7, 99.0%) |
| Tinnitus | 4 | 80/848 (4.7%, 0.6 to 27.5) | < 0.01 (3.9, 97.7%) |
| Sneezing/Coryza | 2 | 290/2768 (8.29%, 3.8 to 17.3) | < 0.01 (0.4, 95.4%) |
| Vertigo | 2 | 11/629 (1.8%, 0.9 to 3.4) | 0.28 (0.0, 13.6%) |
| Otalgia | 1 | 169/2113 (8.0%, 6.9 to 9.2) | NA |
| Epistaxis | 1 | 4/655 (0.6%, 0.2 to 1.6) | NA |
| Hypoacusia | 1 | 1/48 (2.1%, 0.3 to 13.4) | NA |
| Laryngeal sensitivity | 1 | 17/100 (17.0%, 10.8 to 25.7) | NA |
| Nasal itching | 1 | 30/655 (4.6%, 3.2 to 6.5) | NA |
| Cold nose | 1 | 380/2113 (18.0%, 16.4 to 19.7) | NA |
| Phonophobia | 1 | 1/18 (5.6%, 0.8 to 30.7) | NA |
| Dysphagia | 1 | 8/100 (8.0%, 4.1 to 15.2) | NA |
| Voice changes | 1 | 20/100 (20.0%, 13.3 to 29.0) | NA |
| **Respiratory** |  |  |  |
| Dyspnea | 23 | 2829/6275 (34.9%, 23.3 to 48.6) | < 0.01 (1.9, 98.8%) |
| Cough | 17 | 1166/5113 (17.1%, 10.1 to 27.5) | < 0.01 (1.5, 98.0%) |
| Pleuritic pain | 2 | 521/2228 (18.0%, 9.0 to 32.8) | < 0.01 (0.5, 87.6%) |
| Pulmonary fibrosis | 2 | 20/455 (4.5%, 2.9 to 6.9) | 0.6 (0, 0.0%) |
| Xerotrachea | 1 | 423/2113 (20.0%, 18.4 to 21.8) | NA |
| Expectoration | 1 | 6/49 (12.3%, 5.6 to 24.7) | NA |
| Wheezing | 1 | 98/201 (48.8%, 41.9 to 55.6) | NA |
| **Cardiovascular** |  |  |  |
| Chest pain | 13 | 1391/5576 (15.6%, 7.1 to 30.7) | < 0.01 (2.5, 98.9%) |
| Palpitations | 7 | 943/5352 (11.7%, 4.6 to 26.7) | < 0.01 (1.8, 99.1%) |
| Myocarditis | 3 | 21/344 (10.1%, 0.8 to 60.8) | < 0.01 (2.3, 94.9%) |
| Arrhythmia | 2 | 3/318 (1.7%, 0.1 to 23.7) | 0.02 (3.7, 82.7%) |
| Bradycardia | 2 | 4/386 (1.9%, 0.2 to 18.5) | 0.01 (2.6, 83.4%) |
| Pericardial effusion | 1 | 7/26 (26.9%, 13.4 to 46.7) | NA |
| **Gastrointestinal** |  |  |  |
| Diarrhea/Vomiting | 9 | 641/6363 (9.5%, 4.6 to 18.4) | < 0.01 (1.3, 98.5%) |
| Nausea | 4 | 266/2463 (6.7%, 3.4 to 12.9) | < 0.01 (0.4, 72.5%) |
| Dyspepsia | 3 | 23/731 (2.7%, 1.0 to 7.0) | < 0.01 (0.5, 71.3%) |
| Abdominal pain | 1 | 5/49 (10.2%, 4.3 to 22.3) | NA |
| Early satiety | 1 | 1/27 (3.7%, 0.5 to 22.1) | NA |
| **Genitourinary** |  |  |  |
| Urinary urgency | 1 | 1/27 (3.7%, 0.5 to 22.1) | NA |
| Urinary hesitancy | 1 | 1/27 (3.7%, 0.5 to 22.1) | NA |
| **Musculoskeletal** |  |  |  |
| Fatigue/Muscle weakness | 25 | 4872/8520 (46.1%, 32.9 to 59.8) | < 0.01 (1.9, 99.0%) |
| Arthralgia/Myalgia | 13 | 1297/6279 (16.4%, 7.75 to 31.4) | < 0.01 (2.3, 99.2%) |
| Pain and discomfort | 5 | 384/1179 (30.4%, 16.8 to 48.6) | < 0.01 (0.8, 95.9%) |
| Burning feet pain | 2 | 4/382 (2.0%, 0.1 to 45.5%) | < 0.01 (6.5, 90.4%) |
| Numbness | 1 | 1/27 (3.7%, 0.5 to 22.1) | NA |
| Back pain | 1 | 689/2113 (30.0%, 31.1 to 35.7) | NA |
| Sarcopenia | 1 | 11/11 (95.8%, 57.5 to 99.7) | NA |
| **Dermatological** |  |  |  |
| Alopecia | 7 | 685/3622 (17.5%, 9.1 to 30.9) | < 0.01 (0.93, 98.2%) |
| Rash | 4 | 129/2690 (2.3%, 0.6 to 8.3) | NA |
| Allodynia | 1 | 1/27 (3.7%, 0.5 to 22.1) | NA |
| Eczema | 1 | 1/31 (3.2%, 0.5 to 19.6) | NA |
| Hyperhidrosis | 1 | 3/27 (11.1%, 3.6 to 29.3) | NA |
| Hypohidrosis | 1 | 1/27 (3.7%, 0.5 to 22.1) | NA |
| Red spots on toes | 1 | 42/2113 (2.0%, 1.5 to 2.7) | NA |
| **Psychiatric** |  |  |  |
| Insomnia/Sleep disorders | 13 | 1367/5666 (19%, 10.6 to 32.6) | < 0.01 (1.6, 99.0%) |
| Anxiety/Depression | 12 | 835/3861 (24.5%, 16.2 to 35.1) | < 0.01 (0.8, 97.3%) |
| Impaired concentration | 5 | 252/1029 (22.8%, 7.1 to 53.4) | < 0.01 (2.2, 97.5%) |
| Attention deficit | 3 | 26/183 (22.2%, 5.99 to 56.0) | < 0.01 (1.6, 90.6%) |
| Post-traumatic stress | 3 | 133/307 (42.5%, 30.0 to 56.1) | < 0.01 (0.2, 82.0%) |
| Adjustment disorder | 1 | 5/355 (1.4%, 0.6 to 0.3) | NA |
| Obsessive compulsive disorder | 1 | 14/287 (4.9%, 2.9 to 8.1) | NA |
| Panic attack | 1 | 98/734 (13.4%, 11.1 to 16.0) | NA |
| Severe mood swings | 1 | 2/18 (11.1%, 2.8 to 35.2) | NA |

$Tau^2$ is the variance of the effect size parameters across the population of studies, and it reflects the variance of the true effect size (i.e., heterogeneity among studies). $I^2$ refers to the percentage of heterogeneity among the included studies. NA: Not applicable/available; Estimation was done assuming a random effects model.
<sup>a</sup> Results were ordered according to the number of articles included in each meta-analysis.
<sup>b</sup> COVID-19 represents the total of patients reported in selected articles.

### 3.6. Autonomic clusters in post-COVID syndrome

Based on autonomic symptoms (by COMPASS 31) two clusters were obtained (Table 4 – Figure 5A). Impaired visual acuity and blurry vision were more frequently registered during the acute phase in patients belonging to cluster 2 that cluster 1 (Figure 5B), while depression, chills, weakness, diarrhea, musculoskeletal, palpitations/tachycardia, dryness, cognitive involvement, headache, dizziness, and tinnitus were more frequently observed in the post-COVID cluster 2 (Figure 5C). As expected, COMPASS 31 score was higher in cluster 2 than in cluster 1, as were the median Zung scores. Levels of antibodies were not different between clusters (Table 4).

**Figure 5.**
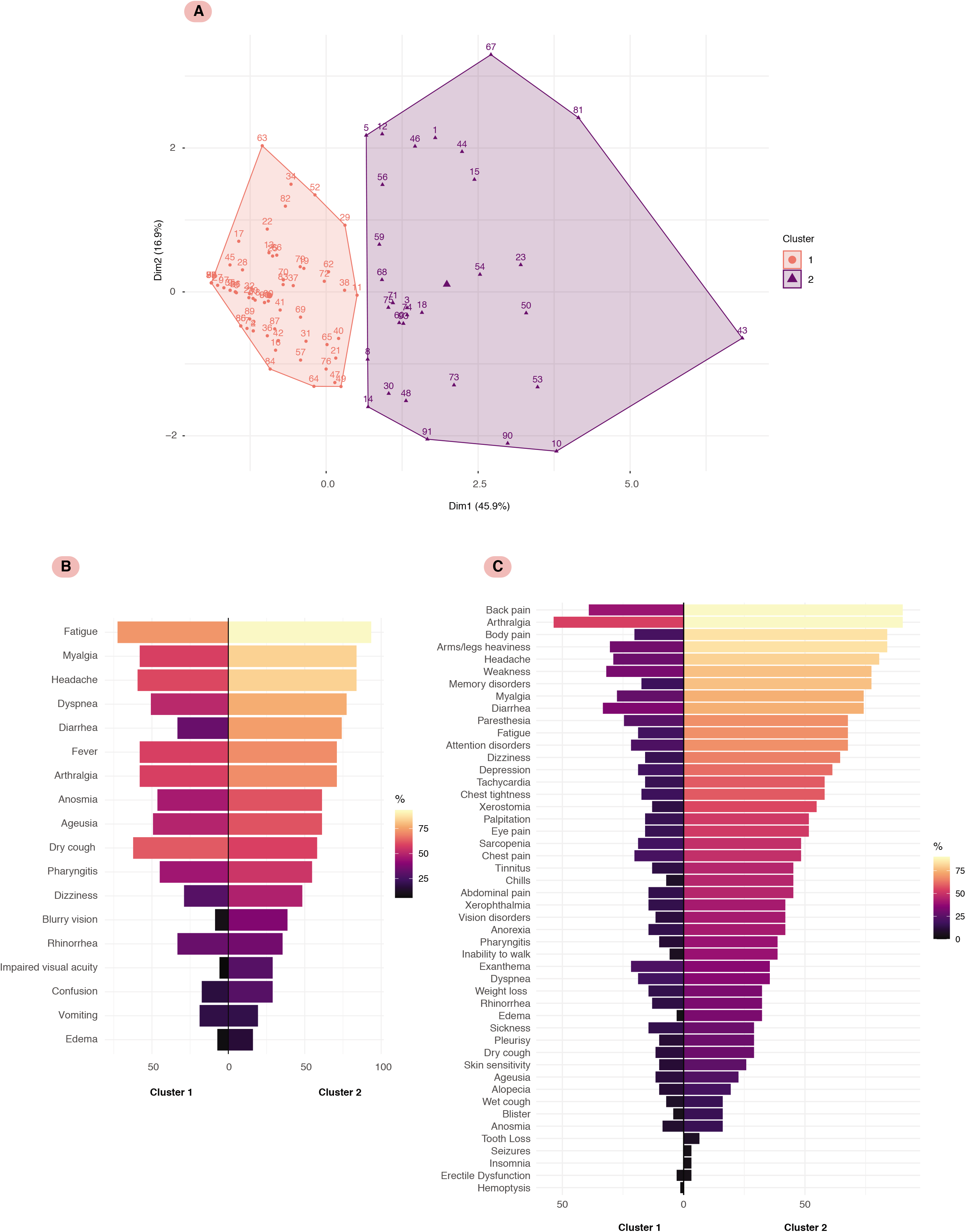
Autonomic clusters in post-COVID syndrome. **A.** Principal components of *K*-means clustering for weighted COMPASS 31 domains. COMPASS 31: composite autonomic symptom score 31. **B.** Mirrored bar plot for acute COVID-19 symptoms on cluster 1 and 2. **C.** Mirrored bar plot for post-COVID clinical manifestations on cluster 1 and 2.

**Table 4.** Cluster analysis based on autonomic symptoms in post-COVID patients.

| Variable | Cluster 1 (n: 69) | Cluster 2 (n: 31) | P value<sup>a</sup> |
| --- | --- | --- | --- |
| **Sex** |  |  | 0.2873 |
| Female | 34 (49.3%) | 19 (61.3%) |  |
| Male | 35 (50.7%) | 12 (38.7%) |  |
| **Age (Median -IQR)** | 50 (41 to 55) | 48 (37 to 55.5) | 0.3748 |
| **Body mass index (Median -IQR)** | 28 (25.2 to 30.2) | 28.1 (25 to 31.4) | 0.9021 |
| **Post-COVID time (days, Median -IQR)** | 202 (111 to 257) | 228 (207 to 257.5) | 0.1177 |
| **Comorbidities (Prior COVID-19)** |  |  |  |
| Hypertension | 12 (17.4%) | 5 (16.1%) | 1.0000 |
| Diabetes type 2 | 10 (14.5%) | 5 (16.1%) | 1.0000 |
| COPD | 1 (1.4%) | 0 (0.0%) | 1.0000 |
| Cancer | 0 (0.0%) | 1 (3.2%) | 0.3100 |
| Dyslipidemia | 9 (13.0%) | 3 (9.7%) | 0.7495 |
| Kidney disease | 2 (2.9%) | 1 (3.2%) | 1.0000 |
| Hypothyroidism | 9 (13.0%) | 3 (9.7%) | 0.7495 |
| Asthma | 0 (0.0%) | 0 (0.0%) | - |
| Coronary artery disease | 0 (0.0%) | 0 (0.0%) | - |
| **Clinical management** |  |  |  |
| Hospitalization | 43 (62.3%) | 22 (71.0%) | 0.4986 |
| Days of hospitalization (Median -IQR) | 11 (8 to 17) | 12 (7.3 to 15.8) | 0.8339 |
| ICU | 15 (34.9%) | 9 (40.9%) | 0.7866 |
| Days of ICU (Median -IQR) | 9.5 (4.5 to 13) | 12 (7 to 15) | 0.4678 |
| **Acute COVID-19 symptoms** |  |  |  |
| Anosmia | 32 (46.4%) | 19 (61.3%) | 0.1981 |
| Ageusia | 34 (49.3%) | 19 (61.3%) | 0.2873 |
| Fatigue | 50 (72.5%) | 29 (93.5%) | 0.0173 |
| Headache | 41 (59.4%) | 26 (83.9%) | 0.0210 |
| Dry cough | 43 (62.3%) | 18 (58.1%) | 0.8249 |
| Rhinorrhea | 23 (33.3%) | 11 (35.5%) | 0.8239 |
| Pharyngitis | 31 (44.9%) | 17 (54.8%) | 0.3931 |
| Dizziness | 20 (29.0%) | 15 (48.4%) | 0.0722 |
| Confusion | 12 (17.4%) | 9 (29.0%) | 0.1960 |
| Impaired visual acuity | 4 (5.8%) | 9 (29.0%) | 0.0028* |
| Blurry vision | 6 (8.7%) | 12 (38.7%) | 0.0006* |
| Dyspnea | 35 (50.7%) | 24 (77.4%) | 0.0155 |
| Diarrhea | 22 (31.9%) | 18 (58.1%) | 0.0163 |
| Vomiting | 13 (18.8%) | 6 (19.4%) | 1.0000 |
| Myalgia | 40 (58.0%) | 26 (83.9%) | 0.0126 |
| Arthralgia | 40 (58.0%) | 22 (71.0%) | 0.2681 |
| Edema | 5 (7.2%) | 5 (16.1%) | 0.2773 |
| Fever | 40 (58.0%) | 22 (71.0%) | 0.2681 |
| **Post-COVID symptoms** |  |  |  |
| Depression | 13 (18.8%) | 19 (61.3%) | <0.0001* |
| New onset AD | 0 (0.0%) | 2 (6.5%) | 0.0939 |
| Anorexia | 10 (14.5%) | 13 (41.9%) | 0.0043 |
| Chills | 5 (7.2%) | 14 (45.2%) | <0.0001* |
| Weight Loss | 10 (14.5%) | 10 (32.3%) | 0.0576 |
| Exanthema | 15 (21.7%) | 11 (35.5%) | 0.2170 |
| Blisters | 3 (4.3%) | 5 (16.1%) | 0.1031 |
| Skin sensitivity | 7 (10.1%) | 8 (25.8%) | 0.0666 |
| Sarcopenia | 13 (18.8%) | 15 (48.4%) | 0.0037 |
| Weakness | 22 (31.9%) | 24 (77.4%) | <0.0001* |
| Diarrhea | 23 (33.3%) | 23 (74.2%) | 0.0002* |
| Myalgia | 19 (27.5%) | 23 (74.2%) | <0.0001* |
| Back pain | 27 (39.1%) | 28 (90.3%) | <0.0001* |
| Arms/legs heaviness | 21 (30.4%) | 26 (83.9%) | <0.0001* |
| Inability to walk | 4 (5.8%) | 12 (38.7%) | <0.0001* |
| Fatigue | 13 (18.8%) | 21 (67.7%) | <0.0001* |
| Body pain | 14 (20.3%) | 26 (83.9%) | <0.0001* |
| Arthralgia | 37 (53.6%) | 28 (90.3%) | 0.0003* |
| Palpitations | 11 (15.9%) | 16 (51.6%) | 0.0005* |
| Xerostomia | 9 (13.0%) | 17 (54.8%) | <0.0001* |
| Chest tightness | 12 (17.4%) | 18 (58.1%) | <0.0001* |
| Tachycardia | 11 (15.9%) | 18 (58.1%) | <0.0001* |
| Edema | 2 (2.9%) | 10 (32.3%) | 0.0001* |
| Chest pain | 14 (20.3%) | 15 (48.4%) | 0.0079 |
| Hemoptysis | 1 (1.4%) | 0 (0.0%) | 1.0000 |
| Dyspnea | 13 (18.8%) | 11 (35.5%) | 0.0818 |
| Wet cough | 5 (7.2%) | 5 (16.1%) | 0.2773 |
| Dry cough | 8 (11.6%) | 9 (29.0%) | 0.0442 |
| Pleurisy | 7 (10.1%) | 9 (29.0%) | 0.0354 |
| Rhinorrhea | 9 (13.0%) | 10 (32.3%) | 0.0301 |
| Sickness | 10 (14.5%) | 9 (29.0%) | 0.1028 |
| Abdominal pain | 10 (14.5%) | 14 (45.2%) | 0.0019 |
| Paresthesia | 17 (24.6%) | 21 (67.7%) | 0.0001* |
| Attention disorders | 15 (21.7%) | 21 (67.7%) | <0.0001* |
| Memory disorders | 12 (17.4%) | 24 (77.4%) | <0.0001* |
| Seizures | 0 (0.0%) | 1 (3.2%) | 0.3100 |
| Headache | 20 (29.0%) | 25 (80.6%) | <0.0001* |
| Dizziness | 11 (15.9%) | 20 (64.5%) | <0.0001* |
| Xerophthalmia | 10 (14.5%) | 13 (41.9%) | 0.0043 |
| Tooth Loss | 0 (0.0%) | 2 (6.5%) | 0.0939 |
| Tinnitus | 9 (13.0%) | 14 (45.2%) | 0.0008* |
| Anosmia | 6 (8.7%) | 5 (16.1%) | 0.3089 |
| Ageusia | 8 (11.6%) | 7 (22.6%) | 0.2243 |
| Vision disorders | 8 (11.6%) | 13 (41.9%) | 0.0011* |
| Eye pain | 11 (15.9%) | 16 (51.6%) | 0.0005* |
| Pharyngitis | 7 (10.1%) | 12 (38.7%) | 0.0017 |
| Alopecia | 7 (10.1%) | 6 (19.4%) | 0.2152 |
| Erectile dysfunction | 2 (2.9%) | 1 (3.2%) | 1.0000 |
| Insomnia | 0 (0.0%) | 1 (3.2%) | 0.3100 |
| **COMPASS 31 score (Median -IQR)** | **6 (3 – 9)** | **22 (18 – 26)** | **<0.0001** |
| **Zung score (Median -IQR)** | **32 (26 – 37)** | **40 (36.5 – 47)** | **<0.0001** |
| **Thyroid stimulating hormone (µIU /mL) (Median -IQR)** | **2.14 (1.4 to 3.2)** | **2.4 (2.0 to 3.5)** | **0.2465** |
| **Total anti-SARS-CoV-2 antibodies (U/mL) (Median -IQR)** | **701.6 (220.7 – 1481)** | **1258 (336.5 – 1945)** | **0.2395** |

IQR: Interquartile range; ICU: Intensive care unit; COPD: Chronic pulmonary obstructive disease; AD: Autoimmune disease; COMPASS 31: Composite Autonomic Symptom Score 31; COVID-19: Coronavirus disease 2019; SARS-CoV-2: Severe acute respiratory syndrome coronavirus 2; Thyroid stimulating hormone, by electroquimioluminescence using the following thresholds: 0.3–4.5 µIU /mL.
<sup>a</sup> P values for categorical variables were obtained by Fisher's exact test. Quantitative variables were analyzed by Mann–Whitney U test.
\* Statistically significant after Bonferroni correction. P value threshold for acute COVID-19 symptoms: 0.05/18= 0.0028; P value threshold for post-COVID symptoms: 0.05/47= 0.0011.

## 4. Discussion

This study indicates that a significant number of patients present with a clinical spectrum after SARS-CoV-2 infection recovery, affecting the quality of life and requiring interdisciplinary approach. Although the cases series do not evaluate incidence nor prevalence, musculoskeletal, digestive (i.e., diarrhea) and neurological symptoms including depression were the most frequent observed in our PCS patients. Arthralgia and diarrhea were the two more frequent acute clinical manifestations persisting during the PCS. Our results were consistent with the meta-analysis in which fatigue/muscle weakness, dyspnea, pain and discomfort, anxiety/depression and impaired concentration were presented in more than 20% of patients. Noteworthy, PCS was independent of severity of acute illness and the humoral response to RBD SARS-CoV-2.

The causes of PCS are under study, however the main hypotheses include a persisting chronic inflammatory process, an autoimmune phenomenon or even a hormonal imbalance as a consequence of an alteration in the hypothalamic-pituitary-adrenal axis [59]. In this line, a study on COVID-19 patients at 3-6 months of convalescence showed that patients with PCS exhibit high levels of CD27^−^ IgD^−^ B cells (which have been associated with autoimmune diseases such as multiple sclerosis [60]), CD8+ T cells, as well as elevated production of Th1 and Th17 cytokines, thus favoring a hyperinflammatory milieu. In addition, patients showed B cell impaired response given by IL-6/IL-10 imbalance [61]. In a similar study, convalescent patients yielded high levels of tumor necrosis factor (TNF) and IL-1β [62]. Recovered patients showed high levels of endothelial activation markers and pro-inflammatory mediators such as growth factors platelet-derived growth factor, vascular endothelial growth factor, MIP-1β, eotaxin, IL-12p70, and IL-17A [63]. Thus, suggesting that clinical manifestations in PCS could be associated to persistence of a pro-inflammatory profile induced by COVID-19 during acute illness. In addition, this may suggest therapeutic targets for the specific management of PCS, including TNF blockade or immunomodulatory drugs.

Based on autonomic symptoms (by COMPASS 31) two clusters were obtained with different clinical characteristics. Cluster 2 exhibited high scores of COMPASS 31. This accounted for the 31% of all patients included (median COMPASS 31 score 22), suggesting that one-third of patients with PCS may yield higher scores when compared with the general population [64]. Interestingly, patients with higher scores exhibited more clinical manifestations and depression. These clusters may have therapeutic implications since clinicians should be aware of particular manifestations during the follow-up, and early psychosocial intervention may reduce the burden of PCS.

Persistence of symptoms has been evaluated in PCS. Moreno-Perez et al. [65], reported that up to 50.9% of the patients considered “recovered” persisted with symptoms similar to those experienced during the acute phase. Moreover, other studies showed that the reported duration of musculoskeletal symptoms varies widely depending on duration of patients follow-up [35,66–68]. Different protocols and populations as well as the preexistence of musculoskeletal symptoms, which were not evaluated in all the studies, preclude a comparative analysis. Nevertheless, early identification would allow to focus the therapeutic management of these patients [69].

Persistent elevation of IL-6 levels, an increase of the angiotensin-converting enzyme 2 in the peripheral nervous system and mast cell participation have been considered to explain the musculoskeletal symptoms in PCS [70–72]. Similarly, over-activation of nociceptive neurons, secondary to neurotropism of the virus, may cause prolongation of symptoms [73]. It is possible that most of the inflammatory response caused by the virus affects the integrity of the central and peripheral nervous system, which promotes the perpetuation of pain after the acute illness [74]. On the other hand, inflammatory damage, triggered by acute infection, would cause psychiatric disorders in predisposed individuals. These symptoms include depression, anxiety and, in worse situations, suicidal behavior [75].

Autonomic dysfunction was confirmed in our study. Orthostatic symptoms and hyperhidrosis have been described [56]. To date, it is unknown if a pre-existing history of minor autonomic symptoms may have some type of relation with autonomic disorders in PCS [76]. These results are relevant since disorders such as postural orthostatic tachycardia syndrome (POTS) and other autonomic dysfunctions may emerge after infectious triggers such as SARS-CoV-2 [77–81].

The mechanisms behind autonomic dysfunction are not clear. However, autoimmune phenomena could be associated with its appearance [82]. Production of autoantibodies against α1, β1 and β2 receptors, angiotensin II receptor type 1, opioids 1, acetylcholine, M2 and M4S receptors are associated with development of POTS [36,83–85]. This immune mechanism, secondary to SARS-CoV2, could be triggered through molecular mimicry between antigens of autonomic nerve fibers, autonomic ganglia, and SARS-CoV2 antigens, as described in some autoimmune conditions.

Respiratory symptoms in PCS are common. The meta-analysis indicated that up to 48% of patients with PCS persisted with respiratory symptoms. Some studies have shown the persistence of pulmonary radiological and functional changes after acute infection [53]. Among the main respiratory functional changes, alteration in gas transfer measured by diffusing capacity for carbon monoxide (D_LCO_) has been documented up to 12 weeks of follow-up [13,86–88]. D_LCO_ alteration could be associated with interstitial and vascular damage, secondary to acute infection [89, 90]. These features may depend on the severity of the disease during the acute COVID-19 [91–93]. Pulmonary radiological alterations have been documented after 12 months post-infection [53].

Some symptoms during acute COVID-19 have been reported to be predictors of PCS, including diarrhea, anosmia, dyspnea, pleurisy, skin sensitivity, and A blood type [15]. A lower SARS-CoV-2 IgG titer at the beginning of the observation period was associated with a higher frequency of PCS [94]. Severity of acute COVID-19 suggests that convalescent critically ill patients commonly experience long-lasting mental health illness. Anxiety, depression, post-traumatic stress disorder, memory and attention disorders are common [95]. However, in our case series severity of acute disease was not associated with PCS.

As expected, the levels of anti-SARS-CoV2 antibodies were lower in patients who were not previously hospitalized [96]. Our results confirm the heterogeneous but long-lasting humoral response to SARS-CoV-2 infection [97, 98]. Other mechanisms beyond the humoral immune response can influence the persistence and/or triggering of PCS, such as viral shedding, therapy, clinical management, immune response, social isolation, comorbidities, age, sex, among others [54]. In terms of viral shedding, some studies have shown that persistent fragments of viral genes, including in feces though not infectious, could induce a hyperimmune response that explaining the persistence of symptoms in post-COVID patients [99, 100]. Moreover, even if the virus is cleared, and there are high neutralizing antibody titers, the immune system could continue to be overactive, thus inducing PCS [99].

## 5. Conclusions

PCS is mainly characterized by musculoskeletal, pulmonary, digestive and neurological involvement including depression. PCS is independent of severity of acute illness and humoral response. Our study confirms the long-term immunity for SARS-CoV-2 but also the inter-individual variability of the immune response. Future studies should evaluate the mechanisms by which SARS-CoV-2 may cause PCS and the best therapeutic options.

## Data Availability

Data will be available prior request to corresponding author

## Abbreviations

CI: Confidence interval.
COMPASS 31: Composite autonomic symptom score 31.
COVID-19: Coronavirus disease 2019.
D_LCO_: Diffusing capacity for carbon monoxide.
ECLIA: Electrochemiluminescence immunoassay.
ELISA: Enzyme-linked immunosorbent assay.
ICU: Intensive care unit.
IQR: Interquartile range.
NA: Not applicable/available.
PCS: Post-COVID syndrome.
POTS: Postural orthostatic tachycardia syndrome.
PRISMA: Preferred Reporting Items for Systematic Reviews and Meta-analyses.
RBD: Receptor binding domain.
S/Co: Signal-to-cut-off.
SARS-CoV-2: Severe acute respiratory syndrome coronavirus 2.
TNF: Tumor necrosis factor.

## Post-COVID study group

Ana María Vargas Suaza, Andrés Mauricio, Palomino Ríos, Carlos Andrés Moya Ortiz, Daniel Fernando Rangel Vera, Diana Carolina Guzmán Núñez, Estefanía Sanabria Medina, Gloria Sofía Guerrero Alvarez, Isabella Casallas Gutiérrez, Isabella Gracia Concha, Isabella Hernández Duarte, Jaime Andrés Antolinez Báez, José Manuel Fernández Rengifo, Jose Manuel Palacio Cardona, Juan Sebastián Beltrán, Julián Francisco Mora Jácome, Laura Zárate Pinzón, María Alejandra Garzón Parra, María Alejandra Melo, Maria Alejandra Muñoz Bernal, María Camila Ayala, María Paula Casanova, María Paula Espitia Correa, Marian Andrea Ochoa Patarroyo, Nicolás Aguirre Correal, Paola Saboya Galindo, Paula Andrea Monje, Santiago Noriega Ramírez, Sara Juliana Guerrero León, Sofía Ballesteros Barreto, Valentina Fragala.

## Acknowledgments

The authors would like to thank all the members of the CREA and Elizabeth T. Cirulli for their contributions and fruitful discussions during the preparation of the manuscript.

