## Supplementary Material 1 for "Post-COVID Syndrome. A Case Series and Comprehensive Review"

Juan-Manuel Anaya^1,2^*, Manuel Rojas ^1^, Martha L. Salinas ^2^, Yhojan Rodríguez^1,2^, Geraldine Roa ^1^, Marcela Lozano^1^, Mónica Rodríguez-Jiménez ^1^, Norma Montoya^2^, Elizabeth Zapata^1^, Post-COVID study group^3^, Diana M Monsalve^1^,

Yeny Acosta-Ampudia^1^, Carolina Ramírez-Santana^1^

^1^ Center for Autoimmune Diseases Research (CREA), School of Medicine and Health Sciences, Universidad del Rosario, Bogotá, Colombia

^2^ Clínica del Occidente, Bogotá, Colombia

^3^ School of Medicine and Health Sciences, Universidad del Rosario, Bogotá, Colombia

***Specificity and Sensibility assessment for Elecsys anti‑SARS‑CoV‑2 S assay***

Elecsys anti‑SARS‑CoV‑2 S assay was tested to evaluate cross-reactivity. 30 samples containing potentially cross-reacting analytes were tested with the Elecsys anti‑SARS‑CoV‑2 S assay (Roche). All the autoimmune diseases, other infectious diseases and pre-pandemic samples were negative by the Elecsys anti‑SARS‑CoV‑2 S assay. No cross-reactivity was found. The resulting overall specificity was 100 % (Supplementary Table 1).

To evaluate sensibility, 16 samples from symptomatic hospitalized patients with a PCR confirmed SARS‑CoV‑2 infection were also tested. All the 16 samples were ≥ 0.8 U/mL in the Elecsys Anti‑SARS‑CoV‑2 S assay and hence considered positive. The median value 2496.5 (IQR, 1644.75 to 3431).

***Correlation between Euroimmun and Elecsys anti‑SARS‑CoV‑2 S assay***

43 samples from acute and convalescent COVID-19 patients, were tested by Euroimmun. The Euroimmun anti-SARS-CoV-2 ELISA (Euroimmun, Luebeck, Germany) was used for serological detection of human IgG antibodies against the SARS-CoV-2 S1 structural protein, in accordance with the manufacturer’s instructions. The ratio interpretation was <0.8 = negative, ≥0.8 to <1.1 = borderline, ≥ 1.1 =positive [1]. The Spearman correlation between these two assays was highly significant (*r =* 0.72; *p =* 2.5 x 10^-8^) (Figure 3A).

***Correlation between neutralizing antibodies and*** ***Elecsys anti‑SARS‑CoV‑2 S assay***

We compared in 18 samples the performance of Elecsys anti-SARS-CoV-2 assay and PRNT_50_, which is the standard for coronavirus serologic analysis. A plaque reduction neutralization test (PRNT_50_) for SARS-CoV-2 was performed using Vero cells, with serum samples diluted from 1:16 to 1:4,096 [1]. The PRNT_50_ correlated with the Roche assay (*r* = 0.52; *p* = 0.00027). (Figure 3B).

***Predictive regression model for neutralizing antibodies at a dilution 1:1024***

We constructed linear regression models with levels of Elecsys anti-SARS-CoV-2 assay as dependent variable to predict PRNT_50_ at different dilutions. These models also included sex, and age as interaction terms. From these models, the model at 1:1024 exhibited the best accuracy for prediction of PRNT_50_ antibodies based on Multiple R^2^ (Supplementary Table 2 –Figure 3C). This model was implemented in a digital platform (<https://crea-covid-19.shinyapps.io/Shiny/>) for its generalized use.

***Estimation of Elecsys anti-SARS-CoV-2 threshold for PRNT_50_ at 1:1024 dilution***

Next, we aimed to estimate an optimal cut-off Elecsys value to detect 50% of SARS-CoV-2 neutralization at PRNT_50_ at 1:1,024 dilution. We used the maximize of the sum of sensitivity and specificity functions to obtain the threshold for 50% of neutralizing activity at this dilution (Figure 3D). This analysis yielded a cut-off value of 151.4 with the maximum sensitivity and specificity achieved, with an area under the curve of 0.78 (Supplementary Table 3).

**Supplementary Table 1.** Specificity of Elecsys anti‑SARS‑CoV‑2 S assay in pre-pandemic individuals.

| Indication | Sample size | Reactive | Specificity (%) |
| --- | --- | --- | --- |
| Autoimmune disease | 10 | 0 | 100 |
| Other infectious diseases (i.e., Zika, Dengue and/or Chikungunya) | 10 | 0 | 100 |
| Pre-pandemic controls | 10 | 0 | 100 |

**Supplementary Table 2.** Accuracy of regression models for prediction of neutralizing antibodies PRNT_50_.

| Dilution | Multiple R^2^ | P value |
| --- | --- | --- |
| 1:16 | 0.1666 | 0.7034 |
| 1:64 | 0.1933 | 0.1210 |
| 1:256 | 0.3582 | 0.0214 |
| 1:1024 | 0.4579 | 0.0348 |
| 1:4096 | 0.3574 | 0.2214 |

**Supplementary Table 3.** Diagnostic accuracy of estimated threshold for 1:1,024

| Cut-off | Sensitivity | Specificity | Positive predictive value | Negative predictive value |
| --- | --- | --- | --- | --- |
| 151.4 | 92.9% | 58.1% | 50% | 94.7% |
